# A Bibliometric Analysis of Preprints in Traditional, Complementary, and Integrative Medicine Research

**DOI:** 10.1101/2025.07.24.25332010

**Authors:** Jeremy Y. Ng, Brenda X. Lin, Sabrina Abdella, Magdalene Abebe, Isabella Y. Tao, Holger Cramer

## Abstract

**Background:** Traditional, complementary, and integrative medicine (TCIM) encompasses a wide range of healthcare practices and is of growing global interest. Preprints, scientific manuscripts posted prior to formal peer review, offer opportunities to address challenges in TCIM research, including limited funding, publication delays, and concerns about methodological quality. By increasing the visibility and speed of research dissemination, preprints may help strengthen the TCIM evidence base. This bibliometric analysis examined the characteristics of TCIM-related preprints posted on servers with TCIM subject filters.

**Methods:** Preprints were sourced from the ASAPbio preprint server directory, limited to servers with TCIM-related categories. Data extraction (June 4, 2024 to January 14, 2025) included title, DOI, abstract, authors, affiliations, preprint posted date, publication journal (if applicable), publication date, preprint type, keywords, number of versions, citations, comments, and funding information. Author geographic distribution, collaboration networks, and key research topics were also analyzed.

**Results:** Between 2012 and 2024, 1,980 TCIM preprints were posted across 11 servers. Research Square hosted the most preprints, and China contributed the highest number. Among the 612 preprints later published in journals, BMC Trials was the most common destination, with a median time of 4.89 months from preprint to publication. Funding information was often missing, but when reported, the National Natural Science Foundation of China was the most frequent sponsor. Overall, citation and comment activity was low. Wellcome Open Research had the highest average citations and comments per preprint among all servers.

**Conclusion:** This study provides the first in-depth analysis of TCIM preprints, revealing active research areas and important gaps in preprint usage, geographical representation, and post-publication engagement. Findings highlight opportunities to improve transparency and research dissemination in TCIM through more consistent preprint practices and tracking.

## Background

### Context and Importance of Traditional, Complementary, and Integrative Medicine

Traditional, complementary, and integrative medicine (TCIM) encompasses a wide array of healthcare practices that fall outside conventional medicine. Traditional medicine is “the sum of the knowledge, skills and practices based on the theories, beliefs and experiences indigenous to different cultures, whether explicable or not, used in the maintenance of health and the prevention, diagnosis, improvement or treatment of physical and mental illness” [1]. Complementary medicine involves “the use of non-mainstream practices alongside conventional medical treatments”, while integrative medicine “brings conventional and complementary approaches together in a coordinated way” [2,3]. The National Center for Complementary and Integrative Health (NCCIH) has three main categories of complementary health approaches: nutritional, psychological, and physical. Nutritional therapies focus on dietary approaches to treatment, such as supplements and herbs. For psychological approaches, therapies such as mindfulness and spiritual practices are included. Lastly, physical treatments encompass approaches such as massage therapy or spinal manipulation. Within these three categories, there are many treatments that intersect two or more classifications such as meditation, which is both a psychological and physical approach [2,4].

TCIM plays an important role in global healthcare, particularly in regions where traditional practices are deeply rooted in cultural heritage. The World Health Organization (WHO) reports that traditional medicine is widely used, with 88% of the global population having utilized some form of TCIM [5]. Patients use TCIM for numerous reasons, such as enhancing quality of life, alleviating symptoms, supplementation to conventional therapies, and gaining control over one’s healthcare [6–8]. This widespread use is paralleled by growing scientific interest and an increasing number of studies exploring the efficacy, safety, and mechanisms of TCIM therapies [9]. The integration of TCIM into modern healthcare systems aims to provide a more holistic approach to health and well-being, addressing the needs of individuals who seek more personalized and culturally resonant healthcare options [2,7].

### Challenges in Traditional, Complementary, and Integrative Medicine Research

The field of TCIM research faces several significant challenges, which have previously been described in detail [10–12]. One of the primary barriers is access to funding [13,14]. Those conducting TCIM research often struggle to secure financial support compared to conventional medical research, limiting the scope and scale of studies conducted [15,16]. High-quality research in TCIM is also hampered by methodological challenges, including difficulties in standardizing treatments and outcomes, controlling for placebo effects, and ensuring rigorous study designs that can withstand scientific scrutiny [17–19].

Additionally, there are competency-related barriers, such as insufficient training and experience in research methods among TCIM practitioners who later seek to research these therapies [13,16]. These issues may contribute to some clinicians and other researchers holding the perception that TCIM research is of relatively lower methodological quality. Bias-related barriers further complicate the landscape of research in this field, with mainstream medical communities sometimes discrediting TCIM practices and research due to personal opinions and anecdotal evidence [20]. This skepticism can lead to the distortion of study results and hinder the publication and dissemination of high-quality studies TCIM research [21].

### Role of Preprints

Preprints are scholarly manuscripts posted on public servers often before formal peer review [22]. Preprints have been gaining traction since the COVID-19 pandemic as a means to rapidly disseminate research findings to the scientific community and the public, providing immediate access to the latest research developments [23,24]. Preprints enhance the visibility and transparency of research by allowing other scientists to read, comment on, and critique the work prior to formal journal publication [25,26]. This informal peer review process can lead to improvements in the manuscript, as authors can receive feedback and consider revisions outside of the peer review process taking place within the journal to which the article was submitted [26]. While some researchers have expressed hesitancy or lack experience in posting preprints, a recent survey has found that many are increasingly using them to disseminate their work more rapidly [27].

In the context of TCIM research, preprints can address several of the aforementioned barriers. By facilitating quicker dissemination and broader access to research findings, preprints can help overcome funding-related access issues (e.g., inability to pay publisher article processing fees) [28]. The open access nature of preprints allows for greater scrutiny and input from a diverse audience, potentially improving the methodological quality of TCIM research [26]. Additionally, the transparency of preprints can help counteract biases by making the entire research manuscript visible, from initial draft to final publication [29]. This increased openness and the potential for early feedback make preprints a valuable tool for potentially advancing the credibility and reliability of TCIM research.

As interest in TCIM and preprinting practices increase, it is crucial to assess the type of TCIM research that are being posted on preprint servers. A research method known as a bibliometric analysis involves the statistical assessment of a subset of publications within a given field and can be used to facilitate an improved understanding of their characteristics and impact [30–33]. Although bibliometric analyses of peer-reviewed publications in the field of TCIM have been performed [34–36], in addition to a scoping review of all TCIM-related bibliometric analyses [37] to date, to our knowledge, no studies have comprehensively explored the characteristics of TCIM preprints. Thus, the objective of this study is to conduct a bibliometric analysis which identifies the quantity and characteristics of preprints posted on the topic of TCIM research.

## Methods

### Open Science Statement

This study’s protocol was registered on the Open Science Framework (OSF) at the following link [38]. The complete set of extracted data used in this bibliometric analysis has been made available on OSF and can be found here: https://osf.io/35nqj. The final manuscript will be preprinted prior to being submitted to a peer reviewed journal.

### Research Question

What are the quantity and characteristics of preprints posted on the topic of TCIM research?

### Study Design

We conducted a bibliometric analysis on TCIM preprints. Bibliometric analysis involves quantitatively evaluating academic literature to discern patterns, impacts, and trends within a particular field.

### Data Sources

Preprints were sourced from select preprint servers listed on the ASAPbio preprint server directory [39].

### Time Frame

The analysis covered all TCIM preprints posted from each included preprint server, with data extracted between June 4, 2024 and January 14, 2025.

### Search Strategy and Inclusion and Exclusion Criteria

Preprint servers on the ASAPbio preprint directory were screened to check whether they had subject area filters for TCIM-related categories (e.g., traditional medicine, complementary medicine, integrative medicine, alternative medicine, etc). The preprint servers were screened, independently and in duplicate, in accordance with whether they included one or more search filters that contained TCIM-related subject area filters. Our search strategy only involved the inclusion of all preprints found across any preprint servers with TCIM-related subject areas to minimize the arbitrary selection of TCIM preprints. All eligible TCIM preprints of any language regardless of whether they belong to additional subject area filters were eligible for data extraction. As such, TCIM preprints posted on preprint servers without TCIM subject area filters, in addition to non-TCIM related preprints on eligible servers were excluded from this bibliometric analysis. This decision was made for practical reasons, due to the challenging nature of manually categorizing preprint topics as belonging to TCIM [4].

### Data Extraction and Analysis

The following data was extracted from each eligible preprint server (as stated by the ASAPbio preprint server directory): preprint server and URLs, preprint server’s disciplinary scope, ownership type, screening processes, external content indexing, permanence of content, preservation of content, commenting. in addition, we also collected the number of yielded TCIM preprints, and what TCIM-related term each preprint server used for their search filer (e.g., traditional medicine, complementary medicine, integrative medicine, alternative medicine, etc.). The following data was extracted from each eligible preprint: preprint server, title of preprint, DOI, preprint posted date (for all versions), status of preprint (e.g., under review in a journal, published in a journal, etc.), journal of final publication (if applicable), final publication date (if applicable), type of preprint (e.g., primary research, review, etc.), abstract of preprint, authors, author affiliation(s), country of corresponding author, funders, number of comments, number of citations, number of versions, and keywords. Microsoft Excel was used to both collect data and then generate descriptive statistics summarizing the bibliometric characteristics of TCIM preprints, including counts, percentages, means, and medians. These statistics were used to identify trends in the collected data.

## Results

In total, 11 preprint servers on the ASAPbio preprint server directory were found to have TCIM subject area filters. **Table 1** includes the full details of preprint server names and their preprint coverage periods. Within these servers, from 2012 to 2024, a total 1980 preprints were extracted, authored by 11 935 unique authors. Of the 1980 preprints, 612 were published in 100 peer-reviewed journals, while the remaining 1368 preprints fell into distinct preprint statuses as portrayed in **Table 2**. Research Square (n=1185) had the greatest number of posted preprints, followed by SSRN (n=480), and OSF Preprints (n=191). There was an average of 1.30 version per preprint, shown in **Figure 1**. 2021 had the greatest number of posted preprints (n=664, 33.56%), followed by 2023 (n=333, 16.83%). Authors were most commonly affiliated with Shanghai University of Traditional Chinese Medicine (n=66, 1.15%), and Beijing University of Chinese Medicine (n=57, 1.00%). The country with the greatest number of preprints was China (n=785, 39.41%), followed by USA (n=143, 7.18%) and India (n=98, 4.92%). Of the published preprints, the most common journal was BMC Trials (n=441, 67.43%), F1000 Research (n=22, 3.36%), and Heliyon (n=21, 3.21%). For journal articles that have been published, between their preprint posting date and final publication date, there was an average of 4.89 months before a preprint was published. Among the preprints, funding information was most often not reported (n=386, 11.36%) or preprints explicitly reported no funders (n=260, 7.65%). However, among preprints that did report funding, the most common sponsor was the National Natural Science Foundation of China (n=277, 8.15%). The full list of posted preprint characteristics can be found in **Table 3**.

**Table 1:** General Characteristics of Preprint Servers with TCIM Subject Filters.

| **Preprint Server Name** | **Preprint Server Coverage Period** | **TCIM Subject Filter(s)** |
| --- | --- | --- |
| **AAS Open Research** | 2018-present | Subject Area: Medical and Health Sciences, Refine Subject Area: Clinical Medicine, Refine Subject Area: Traditional Medicine |
| **Authorea** | 2017-present | Related Keyword: Traditional and Complementary Medicine, Complementary and Alternative Medicine |
| **F1000 Research** | 2012-present | Subject Area: Medical and Health Sciences, Refine Subject Area: Clinical Medicine, Refine Subject Area: Traditional Medicine |
| **Gates Open Research** | 2017-present | Subject Area: Medical and Health Sciences, Refine Subject Area: Clinical Medicine, Refine Subject Area: Traditional Medicine |
| **HRB Open Research** | 2018-present | Subject Area: Medical and Health Sciences, Refine Subject Area: Clinical Medicine, Refine Subject Area: Traditional Medicine |
| **JMIR Preprints** | 2020-present | Subject Area: Complementary and Alternative Medicine |
| **OSF Preprints** | 2016-present | Subject Area: Medicine and Health Sciences, Refine Subject Area: Alternative and Complementary Medicine; Integrative Medicine |
| **Preprints.org** | 2020-present | Subject Area: Medicine and Pharmacology, Refine Subject Area: Complementary and Alternative Medicine |
| **Research Square** | 2018-present | Subject Area: Integrative and Complementary Medicine |
| **SSRN** | 2016-present | Subject Area: Health Sciences, Refine Subject Area: Medical Research Network, Refine Subject Area: MedRN Subject Matter eJournals, Refine Subject Area: Traditional, Alternative, & Complementary Medicines eJournal |
| **Wellcome Open Research** | 2017-present | Subject Area: Medical and Health Sciences, Refine Subject Area: Clinical Medicine, Refine Subject Area: Traditional Medicine |

**Table 2:** Various Categories of Preprint Status Across Preprint Servers with TCIM Subject Filters.

| **Preprint Status** | **Number of Preprints** |
| --- | --- |
| Approved | 31 |
| Published | 612 |
| Revised | 12 |
| Under Revision | 11 |
| Under Review | 257 |
| Posted | 521 |
| Other | 13 |
| Not Reported | 522 |

**Figure 1:**
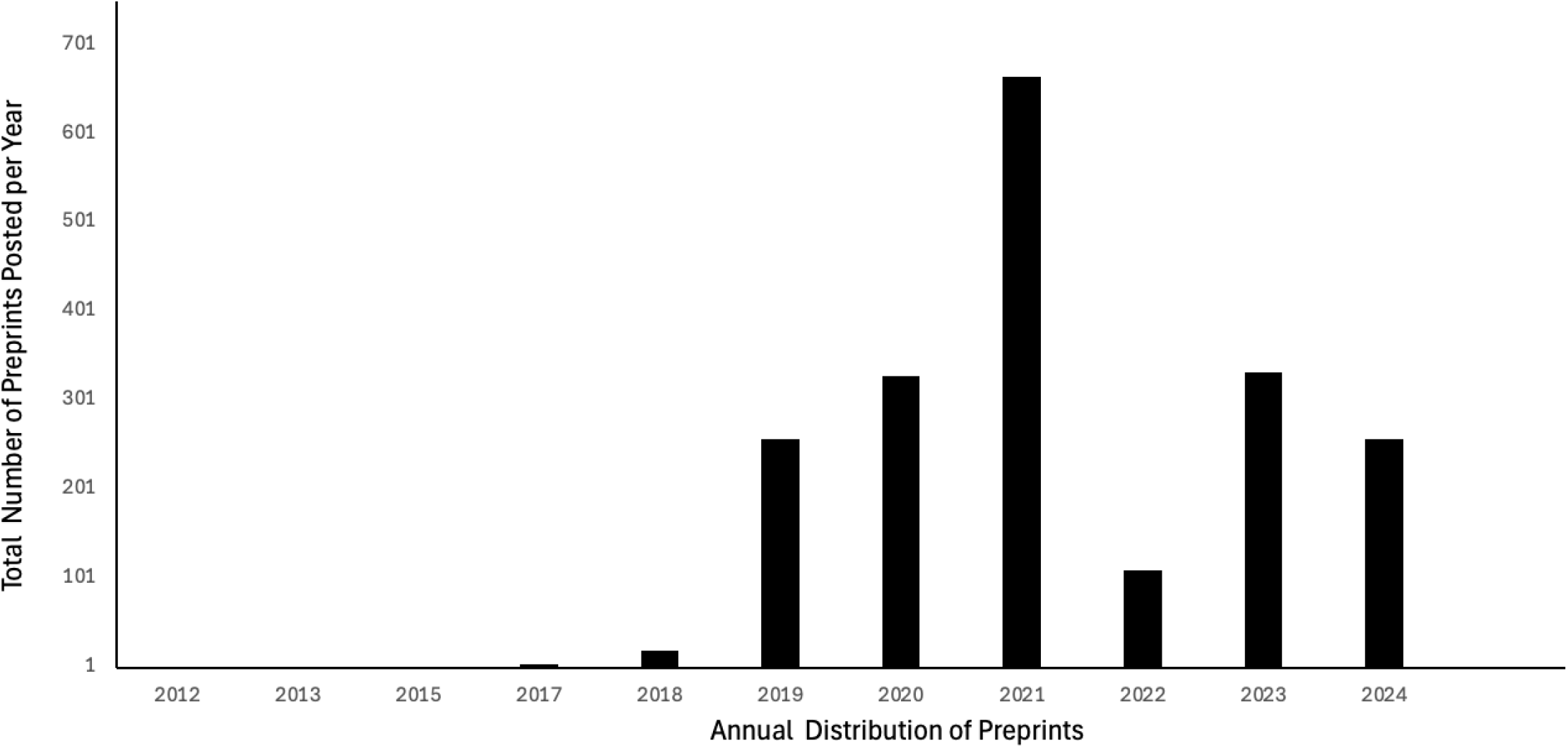
Total Number of Preprints Posted Each Year from 2012 to 2024. **Annual distribution of preprints posted between the years 2012 and 2024.** A total of *n* = 1978 preprints were analyzed using data obtained from the selected ASAPbio preprint server directory. Out of the total 1,980 preprints collected, 2 were withdrawn and lacked associated dates, resulting in 1,978 preprints included in the final analysis. The number of preprints posted per year was as follows: *n=1* (2012), *n* = 2 (2013), *n* = 1 (2015), *n* = 5 (2017), *n* = 20 (2018), *n* = 257 (2019), *n* = 328 (2020), *n* = 664 (2021), *n* = 110 (2022), *n* = 333 (2023), and *n* = 257 (2024). The bar graph illustrates that 2021 had the highest number of preprints posted, whereas the earlier years, 2012 to 2018, had the fewest. In contrast, preprint counts in 2019 and 2020 were comparable to those in 2023 and 2024, while 2022 experienced a notable decline relative to the 2021 peak. Overall, the data revealed a sharp rise in preprint activity beginning in 2019, which peaked in 2021, followed by a significant decline in 2022 and a gradual incline in 2023 and 2024. Conversely, preprint counts between 2012 and 2018 remained consistently low.

**Table 3:**
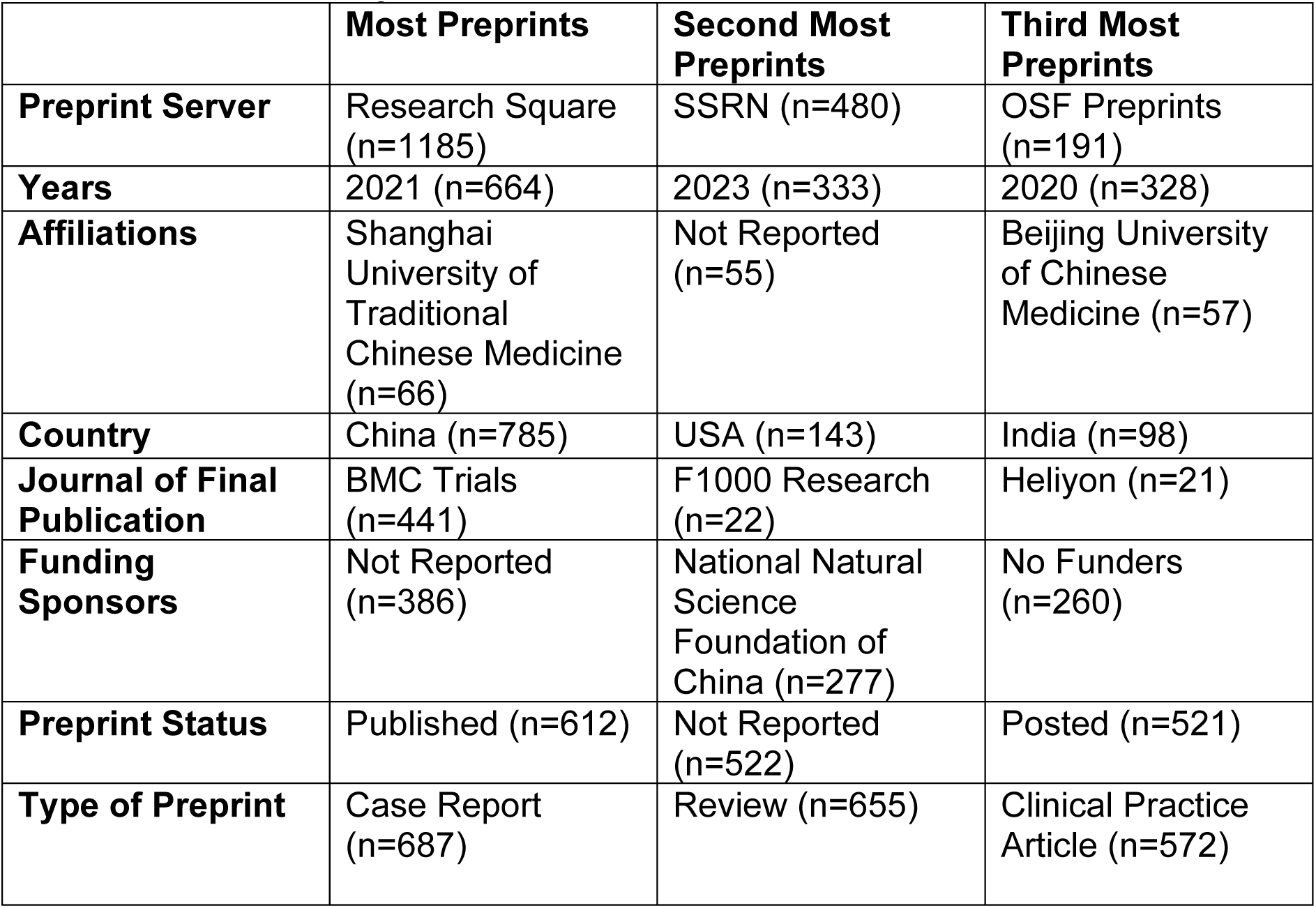
General Characteristics of Posted Preprints on Included Preprint Servers with TCIM Subject Filters.

Overall, there was a low average number of comments (n=0.32) with many not being reported and citations (n=0.83) across all preprint servers. Comments refer to public responses or feedback left by individuals engaging with the preprints on the respective preprint server platforms. The servers with the highest average number of comments were Wellcome Open Research (n=1.75, 90.36%) followed by Preprints.org (n=0.06, 2.99%). The servers with the highest average number of citations across TCIM preprints are Wellcome Open Research (n=8.63, 70.18%) and Research Square (n=1.15, 9.39%). The full list of average comments and citations overall and by journal can be found in **Table 4**. Furthermore, the top 100 keywords appearing amongst the 1980 TCIM preprints were presented according to their frequency of occurrence in **Figure 2**.

**Table 4:** Preprint Comments and Citations by Preprint Server.

| **Preprint Server** | **Average Number of Comments Per Preprint** | **Average Number of Citations Per Preprint** |
| --- | --- | --- |
| **AAS Open Research** | 0 | 2 |
| **Authorea** | Not Reported | Not Reported |
| **F1000 Research** | 0 | Not Reported |
| **Gates Open Research** | 0 | Not Reported |
| **JMIR** | Not Reported | 0 |
| **OSF Preprints** | Not Reported | Not Reported |
| **Preprints.org** | 0 | 0 |
| **Research Square** | 0 | 1 |
| **SSRN** | Not Reported | 0 |
| **Wellcome Open Research** | 2 | 9 |

**Figure 2:**
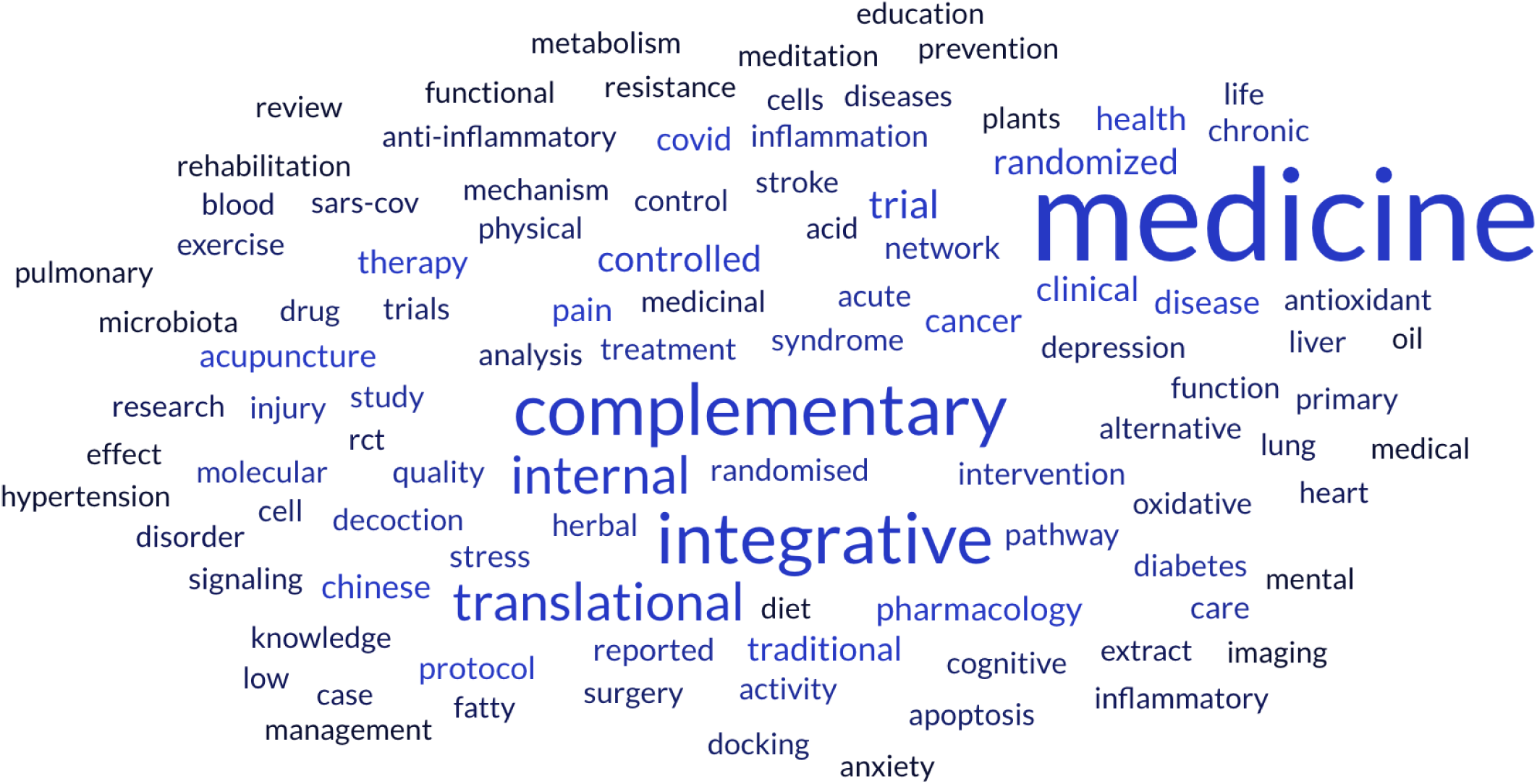
Most Commonly Used Keywords in TCIM Preprints. **Word cloud of the 100 most frequently occurring keywords in preprints published in 1980 across multiple servers.** The size of each word reflects its relative frequency in the dataset. Word cloud generated using *Free Word Cloud Generator* (Source: https://www.freewordcloudgenerator.com).

## Discussion

The objective of this study was to conduct a bibliometric analysis to identify the quantity and characteristics of preprints posted on the topic of TCIM research. To our knowledge, only a few bibliometric analyses have been conducted on preprints, with only two examples within the health/medicine field to our knowledge; these include a bibliometric analysis of preprints on dentistry topics [40] and COVID-19 [41]. The limited number of bibliometric analyses of preprints, when compared to that of published journal articles, is likely attributable to challenges associated with aggregating and processing bibliometric data from multiple preprint servers, which often lack standardized formats. Additionally, standard bibliometric tools used for analysing peer-reviewed publications (e.g., Scopus, Web of Science) may not be compatible with preprint servers.

Over the examined period, TCIM preprints revealed that Research Square was the most frequently used server for posting preprints, followed by SSRN and OSF. This period also includes a notable peak of posted preprints in 2021, as shown in Figure 1, which may be attributed to pandemic related factors. Researchers may have had more time to write and disseminate their findings, potentially contributing to the increased numbers of posted preprints. The prominence of Research Square may reflect its integration with journal submission requirements, accessibility, or greater visibility within the TCIM research community. Its widespread use suggests that researchers may perceive it as a more effective server for reaching relevant audiences or expediting the publication process. In contrast, the lower usage of other servers may be attributed to differences in servers’ accessibility, disciplinary focus, or institutional preferences. These variations highlight the need to explore further the factors influencing researchers’ choice of preprint server, as well as the potential implications for the visibility and dissemination of TCIM research across diverse preprint servers.

Although Research Square hosted the most TCIM preprints, its high volume of preprints did not correspond to greater visibility, highlighting potential challenges associated with preprint oversaturation. Despite its use dominance, Research Square did not rank highest in average citations or comments per preprint. This finding suggests that its large volume of preprints may lead to oversaturation, therefore limiting the visibility of TCIM preprints. In contrast, the preprint server, Wellcome Open Research, which hosted fewer preprints, was found to have a higher number of average of citations and comments per posted preprint. This pattern indicates that authors who choose preprint servers that host fewer TCIM preprints may facilitate greater visibility and engagement per TCIM preprint. In contrast, the oversaturation of TCIM preprints on a widely used server such as Research Square, may dilute attention across a vast number of TCIM preprints internally, ultimately impacting the discoverability of individual TCIM preprints. These variations highlight that the popularity of a preprint server within the realm of TCIM, as measured by the number of TCIM preprints it hosts, does not necessarily translate to high engagement.

An analysis of TCIM preprint status demonstrated that most preprints fell into one of three categories: published as a peer reviewed journal article, not reported, and posted as a preprint. These classifications were used to highlight the importance of tracking the current stage in the research process of TCIM preprints. These findings suggest that, while a large number of TCIM preprints successfully undergo the peer review process and achieve publication, many still remain unpublished in scholarly journals. Given that these statuses are not reported using the same terminology across different preprint servers, this highlights the need for greater transparency and consistency in tracking the progress of preprints to provide a clearer understanding of their roles in the publication process. The remaining preprints were classified into additional status categories: approved for publication in a peer reviewed journal, revised based on peer review feedback, under peer-revision, and other. Other refers to preprints that did not fit the criteria of the categories established such as opinion articles, peer-review reports, short reports, commentaries and theses. This indicates that preprints classified as approved satisfied peer reviewers while those categorized as revised and under revision are in the process of receiving or addressing peer review feedback. The disparity between the high number of posted and unreported preprints and the relatively low number of revised and under-revision preprints emphasizes the need for improved tracking of TCIM preprints throughout the publication process. The data suggests the need for greater measures to ensure preprints receive transparency and attention needed, ultimately increasing publication within TCIM research.

The average number of versions of preprints suggests that most are not subsequently revised or updated, potentially limiting the progression of TCIM research. This pattern reflects the challenges many authors face in advancing their preprints beyond the initial stage. The substantial time, effort, and resources required to revise manuscripts and address peer review feedback can act as significant barriers. As a result, many preprints remain in their initial posted form, highlighting the need for improved support systems that facilitate the revision and publication process. Ensuring that TCIM preprints undergo timely and effective peer review processes, while maintaining the integrity and quality of the work, is essential for strengthening the scientific contribution within TCIM research.

Among the journals that published TCIM research with corresponding preprints from our sample, BMC Trials was the most frequent destination for final publication, followed by Heliyon and BMC Complementary Medicine and Therapies. The high number of publications in BMC Trials suggests that this journal may play a key role in the dissemination of TCIM research. One reason for this observation is the predominance of trials among published articles, which are generally longer and more involved studies [42]. This may create a greater incentive to preprint their results compared to studies such as reviews, observational studies, or bibliometric analyses. A further reason is due to BMC’s agreement with Research Square, which offers an opt-in option for preprint submission when authors submit a manuscript to a BMC journal. This may help explain why BMC Trials hosted a large number of preprints, as the integrated opt-in feature during manuscript submission encourages authors to share their work and increases the likelihood of preprints being posted on Research Square. This observation prompts further inquiry into the factors influencing authors’ choice of preprint servers for TCIM research. It is presently unknown whether authors may select specific servers based on perceived alignment with their research focus, audience reach, platform visibility, ease through a preprint server’s partnership with the selected journal for submission, or other reasons altogether. Regardless, understanding these motivations is crucial for ensuring transparency, reducing potential publication bias, and promoting equitable dissemination of TCIM research across diverse and high-impact scientific platforms.

TCIM preprints were largely concentrated in China, with institutions such as the Shanghai University of Traditional Chinese Medicine and Beijing University of Chinese Medicine contributing a significant proportion. China emerged as a leading contributor to TCIM preprints, underscoring its dominant role in advancing research within this field. This prominence is also evident in funding patterns, with the National Natural Science Foundation of China emerging as a leading funder of TCIM research across our analysed sample of preprints, which may account for the significant representation of TCIM research in China.

A large portion of preprints were classified as case reports, reviews, and clinical practice articles. These results suggest that TCIM research disseminated via preprint servers is largely treatment-focused, with a strong emphasis on documenting clinical observations and patient outcomes. The prevalence of case reports is consistent with the empirical and practice-based nature of TCIM and reflects their relative ease and speed of preparation, often requiring only a clinician’s time and typically completed within a short period. In contrast, systematic reviews were much less common among the preprints analysed. This underrepresentation is likely due to the considerable time, effort, and resources required to complete a high-quality systematic review, which can take many months. Consequently, case reports may be more commonly shared as preprints simply because they are quicker and more feasible to prepare for rapid dissemination. This distribution highlights how practical considerations, such as time investment and ease of preparation, influence the types of TCIM research posted on preprint servers.

### Future Directions

The findings of this bibliometric analysis can inform future research aimed at investigating key factors influencing the progression of TCIM preprints to peer-reviewed publication. Efforts should be directed toward developing standardized tools and methodologies for preprint tracking to enhance consistency and transparency. This is especially important given the large number of preprints that remain unreported or do not progress beyond the initial posted stage. Additionally, it would be beneficial to explore strategies to promote the representation of TCIM practices across different regions, to address the geographic disproportions in TCIM preprint outputs observed. This offers varying perspectives and unique insights on how health concerns in underrepresented regions are addressed utilizing TCIM approaches, therefore, fostering diversity in research perspectives. Exploring author motivations, for utilizing specific preprint servers, may provide valuable insights into the high concentration of TCIM preprints associated with certain journals and publishing platforms. Moreover, it would also be worthwhile exploring why authors of TCIM research may choose not to upload their work to preprint servers. Lastly, exploring the influence of journal-preprint server partnerships, preprint to publication workflow and server visibility would also provide a greater understanding into the publication patterns observed from the studied TCIM preprints. By addressing these areas, future studies can contribute to a more inclusive, transparent, and globally representative preprints within the realm of TCIM.

### Strengths and Limitations

The present bibliometric analysis captured the characteristics of all preprints posted regardless of publication language, which broadened the scope of included TCIM preprints. Due to the complex challenges associated with defining TCIM at the therapy-level, our inclusion criteria relied on preprints available on specific preprint servers that had TCIM subject area filters. While this removed the risk of us introducing human error into the categorization, it is undoubtedly the case that this also resulted in the exclusion of TCIM preprints posted on preprint servers without a TCIM-related subject area filter.

## Conclusion

This bibliometric analysis explored the quantity and characteristics of TCIM preprints. Our findings contribute insights into the evolving field of TCIM research, highlighting areas of high interest and activity while identifying potential barriers related to visibility and the progression from preprint to peer-reviewed publications. This study has also revealed regional disparities in research contributions and limitations in following the progression from preprints to peer-reviewed publications. To our knowledge, this is the first bibliometric analysis on this topic, and it enhances transparency in preprint tracking while informing future efforts to address limitations in early research dissemination within this field.

## List of Abbreviations

NCCIH: National Center for Complementary and Integrative Health
OSF: Open Science Framework
TCIM: traditional, complementary, and integrative medicine
WHO: World Health Organization

## Declarations

### Ethics Approval and Consent to Participate

This study involved a bibliometric analysis of publicly available research posted on preprint servers; it did not require ethics approval or consent to participate.

### Consent for Publication

All authors consent to this manuscript’s publication.

### Availability of Data and Materials

All relevant materials and data are included in this manuscript or posted on the Open Science Framework: https://doi.org/10.17605/OSF.IO/BWD6F

### Authors’ Contributions

JYN: designed and conceptualized the study, collected and analysed data, drafted the manuscript, and gave final approval of the version to be published.

BXL: collected and analysed data, made critical revisions to the manuscript, and gave final approval of the version to be published.

SA: collected and analysed data, made critical revisions to the manuscript, and gave final approval of the version to be published.

MA: collected and analysed data, made critical revisions to the manuscript, and gave final approval of the version to be published.

IYT: collected and analysed data, made critical revisions to the manuscript, and gave final approval of the version to be published.

HC: analysed data, made critical revisions to the manuscript, and gave final approval of the version to be published.

